# Characterization of Childhood Trauma, Hippocampal Mediation and Cannabis Use in a Large Dataset of Psychosis and Non-Psychosis Individuals

**DOI:** 10.1101/2022.05.09.22274865

**Authors:** Elisabetta C. del Re, Walid Yassin, Victor Zeng, Sarah Keedy, Ney Alliey-Rodriguez, Elena Ivleva, Scott Hill, Nicole Rychagov, Jennifer E. McDowell, Jeffrey R. Bishop, Raquelle Mesholam-Gately, Giovanni Merola, Paulo Lizano, Elliot Gershon, Godfrey Pearlson, John A. Sweeney, Brett Clementz, Carol Tamminga, Matcheri Keshavan

**Author notes:** Corresponding Author: Prof. Matcheri Keshavan, Harvard Medical School MMHC Department of Psychiatry, 75 Fenwood Road Boston, MA 02115.

## Abstract

**Background:** Cannabis use (CA) and childhood trauma (CT) independently increase the risk of earlier psychosis onset; but their interaction in relation to psychosis risk and association with endocannabinoid-receptor rich brain regions, i.e. the hippocampus (HP), remains unclear. The objective was to determine whether lower age of psychosis onset (AgePsyOnset) is associated with CA and CT through mediation by the HP, and genetic risk, as measured by schizophrenia polygene scores (SZ-PGRS).

**Methods:** Cross-sectional, case-control, multicenter sample from 5 metropolitan US regions. Participants (n=1185) included 397 controls not affected by psychosis (HC); 209 participants with bipolar disorder type-1; 279 with schizoaffective disorder; and 300 with schizophrenia (DSM IV-TR). CT was assessed using the Childhood Trauma Questionnaire (CTQ); CA was assessed by self-reports and trained clinical interviewers. Assessment included neuroimaging, symptomatology, cognition and calculation of the SZ polygenic risk score (SZ-PGRS).

**Outcomes:** In survival analysis, low CT and CA are associated with lower AgePsyOnset. At high CT or CA, CT or CA are individually sufficient to affect AgePsyOnset. CT relation with AgePsyOnset is mediated in part by the HP in CA users before AgePsyOnset. CA before AgePsyOnset is associated with higher SZ-PGRS and correlated with younger age at CA usage.

**Interpretation:** CA and CT interact to increase risk when moderate; while severe CT and/or CA abuse/dependence are each sufficient to affect AgePsyOnset, indicating a ceiling effect. Probands with/out CA before AgePsyOnset differ on biological variables, suggesting divergent pathways to psychosis.

**Funding:** MH077945; MH096942; MH096913; MH077862; MH103368; MH096900; MH122759.

**Research in context:** *Evidence before this study:* Cannabis use (CA) and childhood trauma (CT) independently increase the risk of earlier development of psychosis. Scarce evidence exists on the interaction between CA and CT and the neurobiological substrate of their interaction.

*Added value of this study:* Analysis of a large transdiagnostic sample of psychosis probands and controls (N=1288) indicates synergy of CT and CA and small but significant contribution of the posterior hippocampus. Data further indicate existence of two populations of probands with psychosis, those with and those without CA after CT before psychosis onset. CT and CA before psychosis onset interact according to a stepwise increase up to reaching a ceiling effect.

*Implications of all the available evidence:* Clinically, youth with low, medium CT need to be targeted for intervention before CA onset.

## Introduction

Age at psychosis onset (AgePsyOnset), associated symptomatology, and both brain morphology and function are modulated by the interaction of genetics and the environment ^1, 2^. Cannabis use ^3^ (CA) and childhood trauma (CT) individually increase the risk for emerging psychosis,^4^ with some evidence demonstrating an additive effect of CT and CA in increasing risk of psychopathology in adolescence.^5^ CT encompasses diverse childhood adversities and is assessed with self-reported scales such as the Childhood Trauma Questionnaire (CTQ) ^6^ or the Adverse Childhood Events (ACE).^7^ In animal models^8^ as well as in humans, ^9^ CT has been shown to affect the hippocampal (HP) volume, albeit not consistently. The HP is one of the brain regions richest in cannabinoid type 1 (CB1) receptors,^10^ the main central nervous system mediators of endogenous cannabinoids and exogenous CA.^10^ Autoradiography binding studies have shown denser binding of agonists in the dorsal (corresponding to the human posterior HP) than in the ventral rodent HP^10^. In translational studies, the effects of CA on memory and learning have long been attributed to its effects on the HP.^11^

CA use often starts in adolescence,^12^ a time of increasing risk-taking, part of the cognitive maturation process, as shown in animal models^13^ and a crucial period for brain development,^14^ when the brain is vulnerable to environmental insults, such as substance use. The endocannabinoid system itself is refined during adolescence,^14^ which is also the period when first signs of psychosis and associated cognitive decline are most frequently observed.^15^

The interaction of CT and CA on the process of psychosis has been evaluated in relation to symptomatology and cognition;^16^ but data on AgePsyOnset^4^ and mediation of CT and CA by neurobiological substrates is limited.^17^ Studies on the accumulation of different genetic and environmental factors on risk of psychosis have indicated involvement of the basal ganglia.^18^ Abnormalities of white matter derived fractional anisotropy have been shown in individuals affected by psychosis with high load of CT and CA compared to controls and unaffected siblings.^17^ There is some evidence of a combined effect of SZ-PGRS with CT, greater than the sum of each alone.^19^ Three-hits models have been proposed where SZ-PGRS might function as a first hit, CT as a second and CA as a third, leading to earlier AgePsyOnset.^15^

Here, CA and CT associations with HP and AgePsyOnset were approached using a large, deeply phenotyped trans-diagnostic sample from the Bipolar-Schizophrenia Network on Intermediate Phenotypes (BSNIP-2) dataset that encompasses bipolar disorder type-1 (BP), schizoaffective disorder (SAD), and schizophrenia (SZ). The evidence of a common neurobiological substrate across diagnoses is robust (e.g., BSNIP studies:^20^; others, e.g.: ^21^ as well as shared genomic relationships).^22^

Our main hypotheses were that 1) CA and CT are associated with lower AgePsyOnset; 2) HP and CA mediate CT effects on AgePsyOnset; 3) the SZ-PGRS might contribute to refine identification of probands with high versus low CT and CA; 4) Probands with CA before versus after AgePsyOnset might differ in key biological variables of interest. We analyzed the data according to presence/absence of psychosis across a trans-diagnostic psychosis continuum to disentangle CA versus non-CA as well as high versus low CT in relation to AgePsyOnset. Other variables of interest were age of CA initiation (AgeCAOnset) and relationship with symptomatology as well as the SZ-PGRS for each participant. The HP was analyzed according to its left and right anterior and posterior portions as previously we showed differential abnormalities in psychoses according to the dorso-ventral division;^20, see also 23^ and for the higher density of endocannabinoid receptors in rodents’ dorsal HP^10^.

## Methodology

### Subjects

Participants (N=1185), included 397 controls (HC) not affected by psychosis and with no DSM Axis1 disorder or first-degree relative with psychotic illness (HC); 209 participants affected by bipolar disorder-type-1; 279 by schizoaffective disorder; and 300 with schizophrenia diagnosed using the DSM IV-TR criteria (Table 1). Data were from participants recruited from the multi-site B-SNIP 2 consortium across Athens and Augusta, GA; Boston, MA; Chicago, IL; Dallas, TX; Hartford, CT. HCs were recruited alongside probands (PRO). The BSNIP2 collected CT data, not available in BSNIP1 dataset. All protocols described were approved by the respective local IRBs. Detailed inclusion/exclusion criteria and comprehensive symptom and cognition battery are in the Supplementary Material.

**Table I.** Clinical and Demographic Characteristics.

| | HC<br>(n=397) | BP (n=209) | SAD (n=279) | SZ (n=300) | $\chi^2/F$ | p value | Significant group<br>comp. |
| --- | --- | --- | --- | --- | --- | --- | --- |
| Age (mean,<br>SD) | 34.30 (11.98) | 36.11 (11.38) | 39.81 (11.72) | 38.12 (11.91) | 15 | <.001 | SAD-HC***, SZ-HC***, SAD-BP*** |
| Sex (M/F) | 164/233 | 90/119 | 132/147 | 184/116 | 29 | <.001 | SZ-HC***, SZ-BP***,<br>SZ-SAD** |
| CA ever use<br>(No/Yes) | 200/197 | 36/173 | 61/218 | 78/222 | 73 | <.001 | BP-HC***, SAD-HC***, SZ-HC***,<br>SZ-BP* |
| Total<br>Childhood<br>Trauma<br>(mean, SD) | 44.84 (10.09) | 57.08 (17.74) | 64.19 (19.37) | 57.72 (16.85) | 107 | <.001 | BP-HC***, SAD-HC***, SZ-HC***,<br>SAD-BP***, SZ-SAD*** |
| BACS<br>Composite<br>Score<br>(mean, SD) | -0.38 (1.23) | -1.01 (1.29) | -1.58 (1.35) | -1.88 (1.31) | 96 | <.001 | BP-HC***, SAD-HC***, SZ-HC***,<br>SAD-BP***, SZ-BP***, SZ-SAD** |
| PANSS Total | -- | 54.38 (1.3) | 63.97 (1.1) | 62.87 (1.1) | 16.9 | <.001 | SAD-BP***, SZ-BP*** |
| PANSS<br>Positive | -- | 12.78 (.42) | 17.09 (.36) | 16.59 (.35) | 33.82 | <.001 | SAD-BP***, SZ-BP*** |
| PANSS<br>Negative | -- | 28.8 (.43) | 31.88 (.37) | 30.29 (.37) | 15.8 | <.001 | SAD-BP***, SZ-BP*** |
| PANSS<br>General | -- | 28.84 (.65) | 31.88 (.56) | 30.29 (.55) | 6.1 | <.01 | SAD-BP** |
| LaHP (mean,<br>SD) | 1588.96 (196.31) | 1573.70 (225.7) | 1525.72 (205.98) | 1523.38 (200.68) | 9.4 | <.001 | SAD-HC***, SZ-HC***, SAD-BP*,<br>SZ-BP** |
| **RaHP (mean, SD)** | **1640.87 (194.92)** | **1637.30 (237.07)** | **1574.66 (203.42)** | **1575.44 (203.96)** | **11** | **<.001** | **SAD-HC\*\*\*, SZ-HC\*\*\*, SAD-BP\*\*, SZ-BP\*\*** |
| **LpHP (mean, SD)** | **1733.08 (191.34)** | **1694.96 (194.80)** | **1665.82 (195.07)** | **1671.75 (187.17)** | **10** | **<.001** | **BP-HC\*, SAD-HC\*\*\*, SZ-HC\*\*\*** |
| **RpHP (mean, SD)** | **1734.45 (180.67)** | **1720.80 (203.74)** | **1688.79 (203.90)** | **1688.76 (191.46)** | **5.2** | **<.001** | **SAD-HC\*\*, SZ-HC\*\*\*** |

HC, healthy controls; SAD, schizoaffective; SZ, schizophrenia; BP, bipolar; L, left; R, right; aHP, anterior hippocampus; pHP, posterior hippocampus; BACS, Battery for the Assessment of cognition; CA, cannabis; CHT, trauma; PANNS, Positive and negative symptoms scale; CU, Caucasian; AA, African-American; OTH, Others; SD, standard deviation; \*, p<0.05; \*\*, p<0.01; \*\*\*, p<0.001

### Genotypes and Polygenic Risk Scores calculation

Genotypes were assessed from blood DNA at the Broad Institute using Illumina Infinium® Global Screening Array-24 v1.0. (GSA) (Illumina Inc., San Diego, CA, USA), containing 688,032 markers mapped to the human genome GRCh37/hg19 reference build. For details and SZ-PGRS, please see Supplementary Material.

### Imaging

Participants received a 3T-MPRAGE scan based on Alzheimer’s Disease Neuroimaging Initiative (ADNI) protocol.^44^ Scans were quality checked for movement, ghosting, and other artifacts and processed with Freesurfer 7.1 (FS7.1). A computational atlas of the hippocampal formation that retrieves the volumetric hippocampal head (anterior) and body (posterior) of each subfield was used.^24^ Each brain scan was rigorously assessed for quality by at least two independent raters. Brain scans with errors in hippocampal segmentation or in the overall cerebral cortex were not included in further analyses (<2%). Detailed delineation of the HP has been described^20^. Estimated total intracranial volume (eTIV) was extracted from Freesurfer.

### Statistical approach

Statistical analyses were performed using SPSS v28. Demographic information for HC and PRO, was analyzed using univariate analyses (ANCOVA) and χ2/F tests. Outliers were considered as >3 SD from the mean and winsorized. The left, right, anterior and posterior hippocampal volumes were compared using MANOVA. For all analyses, data were corrected for age, sex, race, MRI acquisition sites, and in addition for the HP, the eTIV. For CT analyses, the CTQ total score was utilized. For Survival analyses, the CTQ score was operationalized in four strata or quartiles as: CTQ low score, 31-51; moderate, 51-71; medium, 71-91; high, 91-110. Where we found differences between males and females, we report those in the Results. Probability values were corrected for multiple comparisons using False Discovery Rate (FDR).

## Results

### Participants

Significant differences were present in age, sex, and race. 421 participants had CA before AgePsyOnset; 326, after AgePsyOnset. The mean and SD of the composite BACS score and positive and negative symptoms between groups are summarized in Table I. The majority of PRO were treated with second-generation antipsychotics while few were on first generation. There were significant antipsychotic-dose (CPZ-equivalents) differences between SZ and BP and between SAD and BP (BP CPZ: 348·55±398·1, n=158) SAD (CPZ: 528·94±1010·6, n=248) SZ (CPZ: 672·83+-1143·6, n=282); (*p <* 0·01 for both comparisons). There were no significant correlations between CPZ-equivalents and HP volumes, either in CA or non-CA probands. The SZ-PGRS was significantly higher in PRO compared to HC across all stringency values (Supplementary Figure 2A), and higher in probands with CA before than after AgePsyOnset (Supplementary Figure 2B).

### Anterior and Posterior Hippocampus

Analysis of BSNIP2 data confirms significantly smaller volumes of left and right anterior and posterior volumes in PRO compared to HC as reported in del Re et al., 2021 for an analysis that included BSNIP1 and BSNIP2 (Supplementary Figure 1A).

### Cannabis Use

HC subjects were in the none or recreational type CA, while PRO were represented in all 4 categories: No CA, recreational use, *abuse*, and *dependence*.

AgePsyOnset was significantly lower for CA probands compared to that of non-CA (F=4.24, pFDR =0.039); with no significant difference in AgePsyOnset for CA after or before AgePsyOnset (F=2.17, pFDR= 0.091). The self-reported AgeCAOnset though differed significantly in PRO across usage category (F=5.3; *p*<0.001), where for PRO with a history of CA dependence, AgeCAOnset was significantly lower than PRO with recreational CA (*p*=0.009). AgeCAOnset was significantly and positively correlated with the left and right HP in PRO, i.e., higher HP volume correlated with higher AgeCAOnset (left HP, r=0.11, *p*<0.001 and right HP, r=0.11, *p*<0.001) as well as inversely with CTQ score (r=-0.15, *p*=0.0003), where the higher the trauma, the younger the AgeCAOnset. The relationship between CT and AgeCAOnset usage was driven by probands with CA before AgePsyOnset (r=-0.23; p= 3.4041E-10 versus r=-0.06; p=0.373 for participants with CA after AgePsyOnset). Significant correlation between the SZ-PGRS and AgeCAOnset was found for probands with CA before AgePsyOnset, but not for the whole probands’ group or probands with CA after AgePsyOnset (p>0.05 at all PT stringencies; Data in Supplementary Table 1).

### Childhood Trauma

PRO had higher trauma-CTQ scores than HC, as expected and the total CTQ score was significantly higher in CA versus non-CA PRO (F =4.56; p=0.033). Among users, those with CA after AgePsyOnset had higher CTQ than PRO with CA before AgePsyOnset (67.1± 19.5 vs 59.0 ± 16.5, respectively; F=34.8, p= 5.4207E-9). Nonetheless, only in PRO with CA before AgePsyOnset, the total CTQ score was significantly and negatively correlated with the right anterior (r=-0.11; pFDR0.011) and the left and right posterior hippocampus (r=-0.14; pFDR=0.0004; and r=-0.12; pFDR=0.003; Supplementary Figure 1D), where to a higher CTQ score corresponded lower HP volume. CTQ-HP correlations were not significant in PRO with CA after AgePsyOnset (Supplementary Figure 1C) and HC (Supplementary 1B). In PRO with CA before AgePsyOnset, CTQ was also significantly and negatively correlated with AgePsyOnset (r=-0.221; p=1.5581E-12).

### Symptomatology

The total positive PANSS score was positively correlated with the trauma-CTQ score (r=0.24, p<0.001); this was not found for the total negative PANSS score (r=0.08, p=0.07); the positive PANSS score correlated as well, inversely, with AgePsyOnset (r=-0.125, p=0.005), while the correlation between AgePsyOnset and negative symptoms was not significant (r=-0.03, p=0.495). The positive PANSS score here in the BSNIP2 dataset also inversely correlated with the left, right anterior and left posterior hippocampus (r=-0.095, p<0.001; r=-0.089, p=0.001, and r=-0.092, p=0.001respectively) but not the right posterior hippocampus (r=-0.068, p>0.05). Correlations between PANSS negative score and the HP were not significant. Both negative and positive symptom scores were significantly and inversely correlated with cognition (total BACS score; r=-0.29, p<0.001 and r=-0.156, p<0.001).

### Relationships between trauma, cannabis use, cognition, and the HP

The left and right anterior and posterior HP were positively and significantly correlated with BACS score in the whole PRO sample (r=0.20, p=2.9674E-7; r=0.194, p=6.2953E-7; r= 0.18, p=0.000004; r=0.17, p=0.000013) and in PRO with CA before AgePsyOnset (r=0.24, p=0.000004; r=0.24, p=0.000003; r=0.17, p=0.000886; r=0.15, p=0.004, respectively), but not in CA users after AgePsyOnset (all p>0.05).

### Regression Analysis: Variables that Predict Group

To determine which variables predict Group, we adopted a logistic regression model that included the following variables: left, right, anterior and posterior HP, total trauma-CTQ score, composite BACS score, and use or no use of CA; as well as the SZ-PGRS at p=5 × 10^−8^. We included the left and right paracentral volumes as control regions. The results are in Table II. The B values indicate that CA, higher CT, low BACS score, small left posterior HP and higher SZ-PGRS score (PT 5 × 10^−8^) predict proband status. Control regions and other HP portions, did not contribute to the model and were not included further in analyses.

**Table II.** Regression analysis.

|  | B | S.E. | Wald | df | Sig. | Exp(B) | 95% C.I. for EXP(B) |  |
| --- | --- | --- | --- | --- | --- | --- | --- | --- |
|  |  |  |  |  |  |  | Lower | Upper |
| Left Hippocampus Anterior | -.001 | .001 | .248 | 1 | .618 | .999 | .997 | 1.002 |
| Right Hippocampus Anterior | .001 | .001 | 1.806 | 1 | .179 | 1.001 | .999 | 1.003 |
| Left Hippocampus Posterior | .003 | .001 | 9.099 | 1 | .003 | 1.003 | 1.001 | 1.005 |
| Right Hippocampus Posterior | -.002 | .001 | 2.920 | 1 | .087 | .998 | .996 | 1.000 |
| PGRS (PT 5x10<sup>-8</sup>) | -.264 | .095 | 7.730 | 1 | .005 | .768 | .638 | .925 |
| Cognition-BACS total score | .640 | .080 | 64.485 | 1 | <.001 | 1.897 | 1.622 | 2.217 |
| Childhood Trauma-low/high | 1.638 | .200 | 66.797 | 1 | <.001 | 5.145 | 3.473 | 7.620 |
| Left Paracentral volume | .000 | .000 | 1.007 | 1 | .316 | 1.000 | 1.000 | 1.001 |
| Right Paracentral volume | .000 | .000 | .722 | 1 | .395 | 1.000 | .999 | 1.000 |
| CA use (yes/no) | 1.354 | .196 | 47.886 | 1 | <.001 | 3.874 | 2.640 | 5.685 |
| Constant | -5.277 | 1.125 | 21.992 | 1 | <.001 | .005 |  |  |

All variables were adjusted for site, sex, race, and the hippocampus for intracranial volume as well. CI, confidence intervals

### Interactive effect of Trauma and CA use on age at first symptomatology onset

Kaplan-Meyer Survival analysis was employed to determine the combined effect of CT and CA on AgePsyOnset. We considered the whole cohort, stratified by trauma: low, moderate, medium, and high; and CA use: none, recreational, abuse and dependence. The dependent time-variable was time of first reported symptom. As HC are not psychotic, their data in the analyses were censored (See details Supplementary Table 2). Results, in Figure 1A and B and in Table III, indicate that higher CT is associated with earlier AgePsyOnset in both CA and non-CA individuals. For CA, recreational CA in the presence of low trauma, is sufficient to lower AgePsyOnset, indicating that CA adds on CT to lower AgePsyOnset. Visual inspection of curves indicates that high trauma or otherwise high CA are each sufficient to affect AgePsyOnset. Replication in CA before AgePsyOnset only, produces similar results (Figure 1B).

**Table IIIA, B and C.**
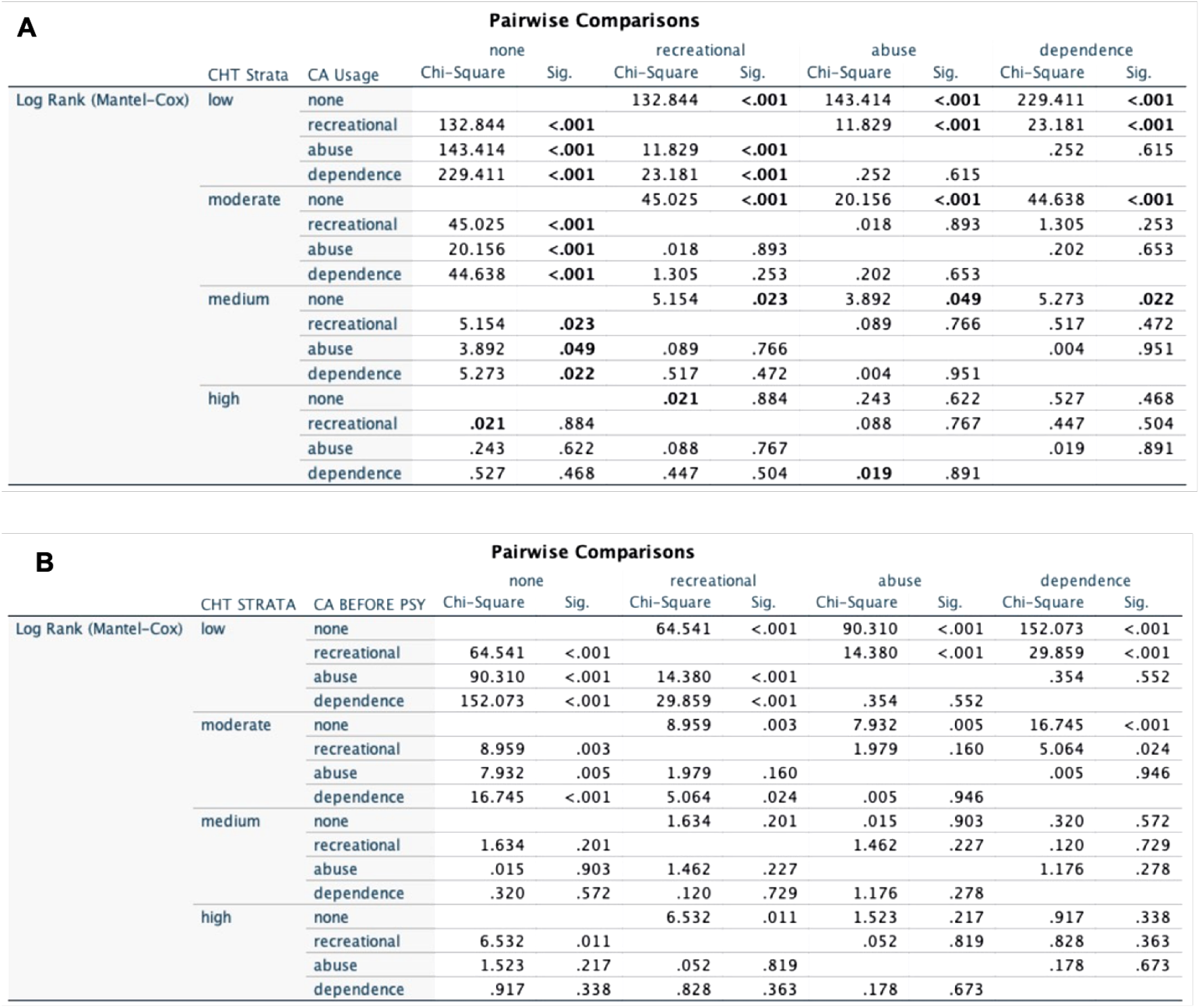
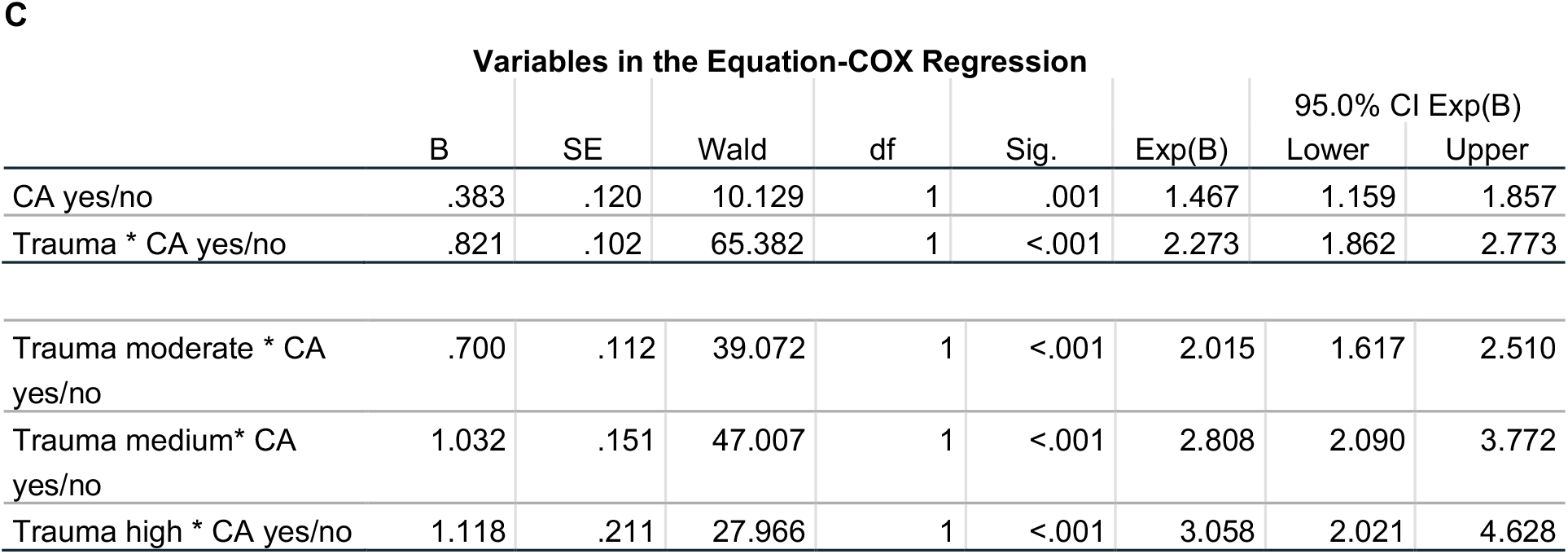
(A) Comparisons of Survival curves by Log Rank Mantel Cox in the whole sample and (B) in CA before AgePsyOnset only. (C) Results of COX Regression analyses.

**Figure 1A and B.**
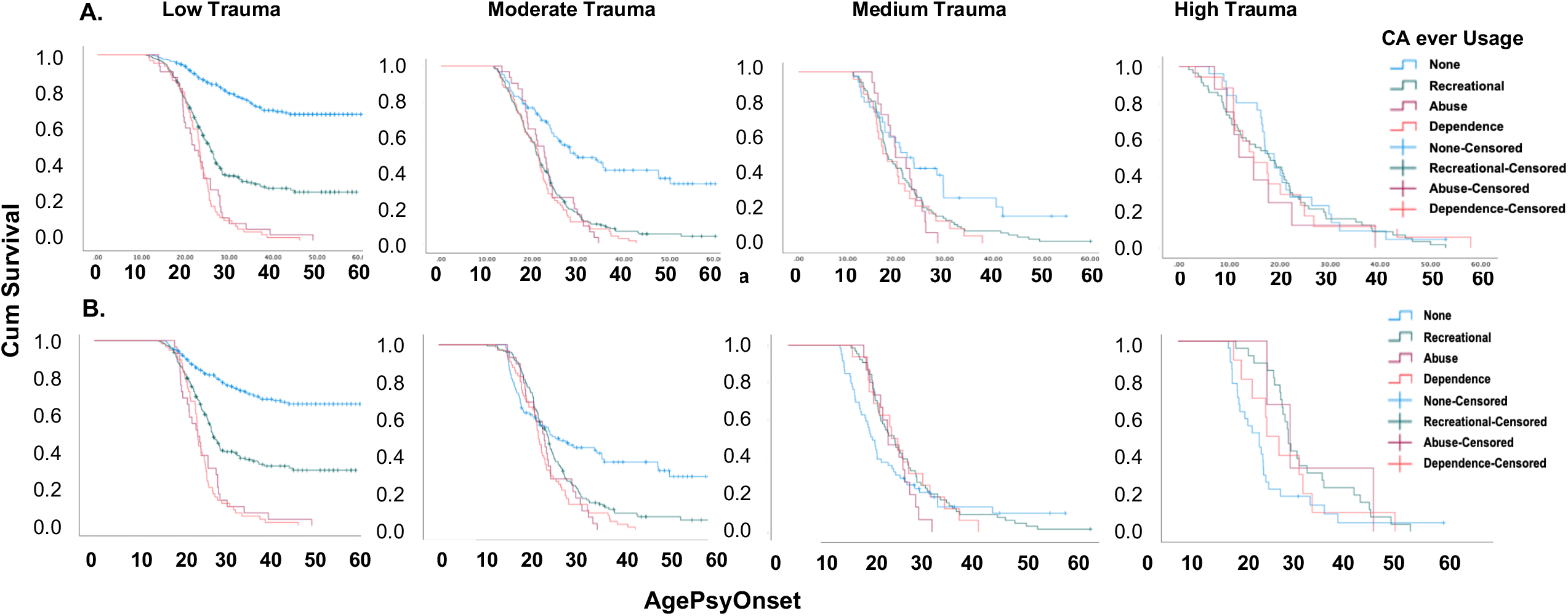
Time to Symptomatology Onset According to Different CA Usage and Timing and Levels of trauma. **A**. CA usage at any time during the lifetime at different levels of trauma and CA usage as indicated. **B**. Here, CA usage is considered only if before AO. trauma, Childhood trauma; CA, Cannabis; Cum, Cumulative; AgePsyOnset, Age at first Symptomatology.

#### *Log-rank (Mantel-Cox)* testing

Overall comparison of survival curves indicates that survival differs significantly for various CA usage at the different trauma levels [χ^2^: 247.9; 3 df (degree of freedom); pFDR<0.001]. Pairwise comparison of curves in Figure 1A and B at different CA across different CT strata are tabulated for the overall sample as well as in CA users before AgePsyOnset (Table IIIA and B).

**Figure 2A and B.**
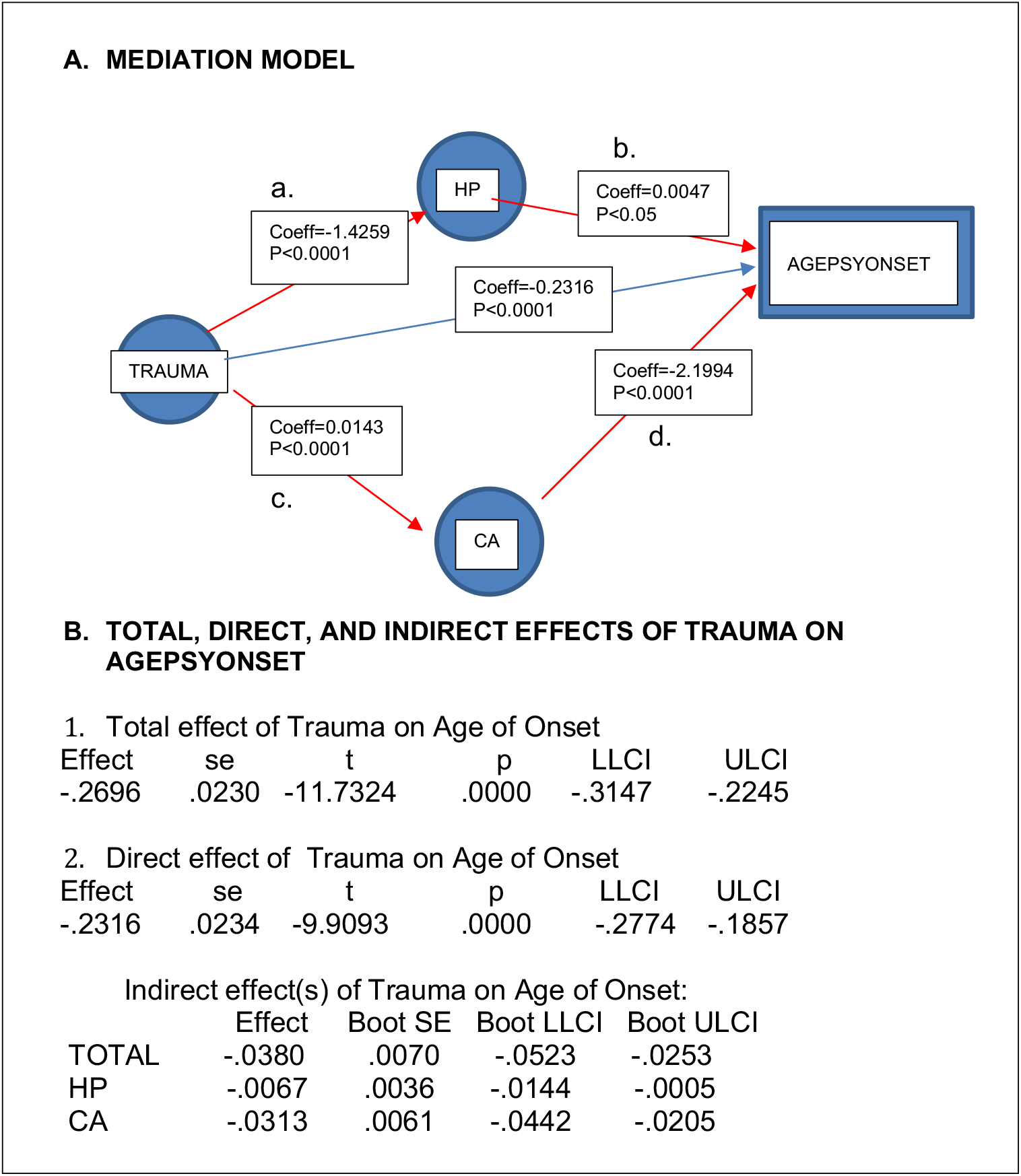
A. Mediation Model. Here the dependent variable is AgePsyOnset. The direct effect of trauma on AgePsyOnset is represented by the blue arrow, while in red are represented the mediation effects of CA and HP. Trauma has a significant effect on the left posterior hippocampus and the hippocampus has a small but significant effect on the age of psychosis onset. Trauma has also a significant effect on CA and CA has a significant effect on age of onset. The direct effect of trauma on age of onset has a significant correlation coefficient=-0.23.16; while the model including CA and HP has a significant coefficient of -0.2696. Thus, the coefficient for the total model is explained by a small but significant contribution of CA and HP. Unstandardized path coefficients are displayed along with standard errors in parentheses, and significance levels (ie, p value) underneath them. Note that the product of paths a and b (a x b) is the indirect effect of Trauma on AgePsyOnset through the hippocampus; while the product of paths c and d (c x d) is the indirect effect of trauma on AgePsyOnset through CA. The total effects, direct effects, and indirect effects were considered significant when the p < 0.05 or 95% CIs did not contain zero. **B. Direct and indirect effects’ coefficients for the model**. Note HP, left posterior hippocampus; CA, cannabis; AGEPSYONSET, age at psychosis onset; Coeff, coefficient; se, standard error; LL CI, lower confidence intervals, ULCI, upper confidence intervals.

### COX Regression analyses

We followed up Kaplan-Meyer and Log-rank (Mantel-Cox) analyses, with a prediction model that considers the time to survival variable, through COX regression modeling. In the model, we considered low versus high trauma, and ever CA, with an interaction term between cannabis ever use and low or high trauma. The results are summarized in Table IIIC. The omnibus test for model coefficients indicates that the overall model is significant (χ^2^=131.354, p<0.001). The Exp(B) values were 1.467 (C.I. 1.159-1.857) for the CA only prediction (p=0.001); and 2.273 (C.I. 1.862-2.773) (p<0.001) for the interaction term, indicating higher hazard ratio for earlier AgePsyOnset when CA and CT are both present. By inspecting the Log-log graph, we determined that curves for the variables did not cross, lending validity to the model.

### Mediation Analysis

To understand the relationship between trauma and HP, we carried out a mediation analysis where we compared the direct effect of trauma on AgePsyOnset and a model that includes the indirect effects of trauma through the mediators CA and left posterior hippocampus (Figure 3). Here, the direct effect of trauma (unstandardized coefficient) on AgePsyOnset is significant and equal to: -0.2316 (se .0234) [t=-9.9, p<0.0001; CI (−0.277, -0.1857)]. When including CA and HP in the model, the coefficient is -0.2696 (se 0.023) [t=-11.73, p<0.0001; CI -0.3147, -0.2245)], indicating significant effect of the mediation variables. The indirect effects contributing to the total coefficient in the model are, CA with a coefficient of -0.0313 and HP with a coefficient of 0.0247. Booth tests reported in Figure 3B confirm that the effects of CA and HP are significantly contributing to the model. The R^2^ for the model including HP and CA as mediator is 0.179, explaining 17.9% of the variance in the model; the R^2^ of the direct trauma effect model is 0.1421, explaining 14% of the variance in the model.

## Discussion

By employing a large, deeply phenotyped transdiagnostic dataset of psychosis probands and non-psychotic controls, we tested the hypothesis of an interactive effect between CT, genetic risk, cognition and CA on AgePsyOnset; and mediation of CT by the hippocampus and CA. The analysis assessed relationships between AgePsyOnset; CA and AgeCAOnset; CTQ total score; Composite cognition score; left/right/anterior/posterior HP and the SZ-PGRS.

In logistic regression, the left posterior HP, CT, CA, cognition and the SZ-PGRS all predict PRO vs no-PRO status. The variables further correlate with AgePsyOnset, where 1. lower cognition, 2. CA, and 3. CT, predict younger AgePsyOnset. To determine the effect of different levels of trauma and CA on survival, i.e. time to AgePsyOnset, we employed Kaplan-Meyer Survival analysis. Here CT and CA interact to increase risk at intermediate CT and CA levels; instead, high levels of CT or otherwise high levels of CA, are individually sufficient to affect AgePsyOnset. Thus evidently, there is a stepwise worse survival with increasing levels of CT and CA, reaching a ceiling effect at higher CT and high CA levels. This is evident in Figure 1 and Table III, where pairwise comparison of survival curves confirms graphic survival analyses.

By use of COX regression analysis, we find that AgePsyOnset is lower with CA, and lower with both CT and CA as shown by CA-CT interaction. Step-wise increase of hazard ratios on AgePsyOnset are included in Table III C. Looking at Di Forti report of odd ratios (not hazard ratios) for symptomatology risk attributable to CA^3^, and Shafer and Fisher’^25^ report of odd ratios for risk with respect to trauma, the hazard ratios we report are similar in their step wise risk increase with increased CA and higher CT. As a novel observation, we show that when combined, CA and CT interact, with an additional risk over that of each variable alone especially at low levels of CT and CA. Harley and colleagues,^5^ in an elegant analysis, had also reported that the sum of CA and CT risk is above that of the individual variables. The limitation of that study was the relatively small sample size. In the current study, we find that the SZ-PGRS contributes to regression models in predicting Group but not survival; further in PRO with CA before AgePsyOnset, the SZ-PGRS significantly correlates to AgeCAOnset. The literature in this regard is mixed, with some reports of a genetic contribution to CA and psychosis,^26^ and possible synergy between SZ-PGRS and CT.^19^ In a study of risk in five major psychiatric disorders, a high correlation between genetic risk for SCZ and cannabis involvement is reported.^27^ Collectively, our analyses can be interpreted as indicating the existence of two probands groups: Probands with CA before and those with CA usage after AgePsyOnset. These two groups are clearly distinguished by the SZ-PGRS (higher in CA before AgePsyOnset); by CT (lower in probands with CA before AgePsyOnset as compared to probands with CA after); And by significant (in probands with CA before AgePsyOnset) versus non-significant correlation between CT, cognition, and the hippocampus. We propose that the neurodevelopmental path of probands characterized by higher SZ-PGRS, lower CT (versus probands CA usage after AgePsyOnset), and CA before AgePsyOnset can be modeled according to a three hits pathway,^15^ while the higher CT load in CA after AgePsyOnset might be sufficient to lead to psychosis without the need of other hits, i.e. CA.

Other variables of interest in the study were cognition that data indicate being lower with CT and CA and predictive of lower AgePsyOnset in line with other studies reporting that cognition, impaired in many mental illnesses^28^, is affected by CT and CA. Data in the literature though lack consistency^29^. For example, a smaller study has shown better cognition in SZ after adolescent CA^30^. Our novel contribution indicates that cognition is inversely related to the hippocampus in probands with CA before AgePsyOnset, but not after, indicating a possible confounder in previous analyses. Our mediation model further indicates that CA and hippocampus mediate a small but significant effect of CT on the AgePsyOnset, providing clues and possible neurobiological therapeutic targets for CT and CA in relation to AgePsyOnset.

There are limitations to our study. The cross-sectional nature of the study is a limitation and will limit causal inferences. CA here is considered without knowledge of its composition, as ‘miscellaneous CA’. Another limitation is that the AgePsyOnset and trauma are self-reported as in the majority of studies in the literature. Nonetheless, the recent study by Danese^31^ indicates that the subjective experience of childhood maltreatment in psychopathology is meaningful compared to objective, clinical measures. Further, childhood trauma questionnaire (CTQ) correlations have been found to show reasonable correlations with prospective violence assessment in children and adolescents.^32^ The research on this by Newbury (2018) indicates that retrospective self-report of childhood maltreatment has strong association with psychiatric problems, lending validity to retrospective analyses. In this context, to add validity to the data, the BSNIP raters were asked to rate confidence in self-reporting after each participant’s interviews. On a scale of 1 to 3, the mean confidence value was 1.29 with a median of 1.0. In subdividing groups of PRO in CA before or after AGEPSYONSET; and with different levels of trauma and CA, groups might be small with less power.

CA is one of the substances most widely used in the world. Recently, the American Psychiatric Association released recommendations that CA should be strictly regulated, including limitations in marketing to children and adolescents (see also ^33^).

All in all, we propose that there are two populations at risk for early AgePsyOnset, characterized by different levels of biological variables and environmental risks which point to the necessity of focusing therapeutic interventions not only on extreme cases of CT and CA, but on adolescent groups were CT and CA are at intermediate levels and where CA*trauma synergy is active.

## Data Availability

All data produced in the present study are available upon reasonable request to the authors

## Acknowledgements

We thank all the participants that generously participated in the study. We also thank Dr. Roger Davis, for statistical consulting. Dr. Davis was supported by the Harvard Catalyst. The Harvard Clinical and Translational Science Center (National Center for Research Resources and the National Center for Advancing Translational Sciences, National Institutes of Health Award UL1 TR002541 and financial contributions from Harvard University and its affiliated academic health care centers. The content is solely the responsibility of the authors and does not necessarily represent the official views of Harvard Catalyst, Harvard University and its affiliated academic health care centers, or the National Institutes of Health.

## Supplementary Material

**Supplementary Figure 1A, B, C, D.**
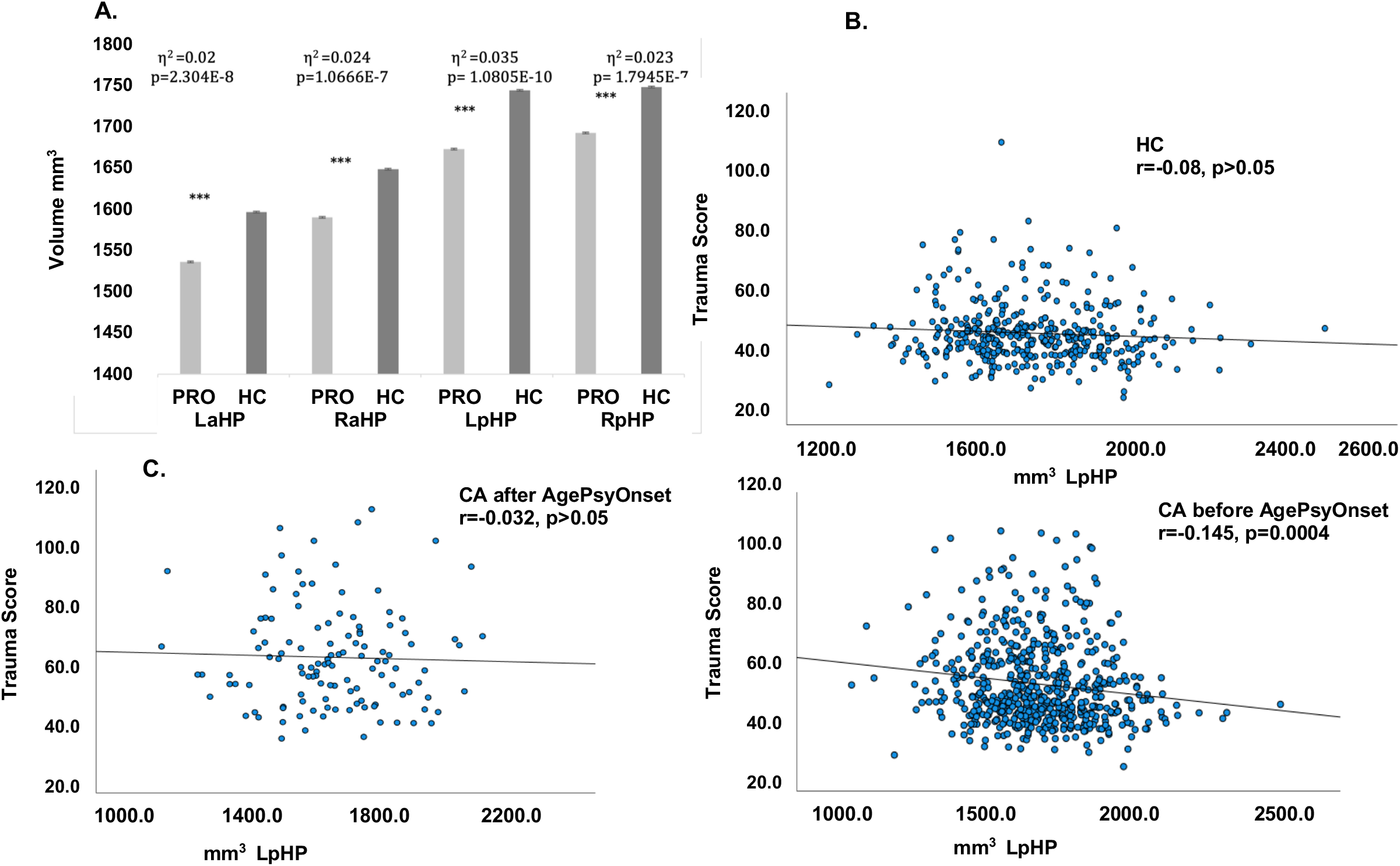
**A**. Volumetric differences of hippocampal volumes between probands (PRO) and controls (HC), whiskers represent standard deviations. (Left anterior HP [F(1,834)=39.16; p<0.001]; Right anterior HP [F(1,834)=29.35; p<0.001]; Left posterior HP [F(1,834)=28.35; p<0.001]; Right posterior HP [F(1,834)=18.97; p<0.001). **B**. Correlation between LpHP and total trauma is not significant in HC; **C**. Correlation between LpHP and Total trauma is not significant in PRO with CA usage after AgePsyOnset; **D**. Significant correlation between LpHP and total trauma in PRO with CA usage before AgePsyOnset. LaHP, left anterior hippocampus; RaHP, right anterior hippocampus; LpHP, left posterior hippocampus; RpHP, right posterior hippocampus; Trauma, Childhood trauma measured by the total CTQ score; CA, cannabis; η^2^, effect size for MANOVA analysis

**Supplementary Figure 2A and B.**
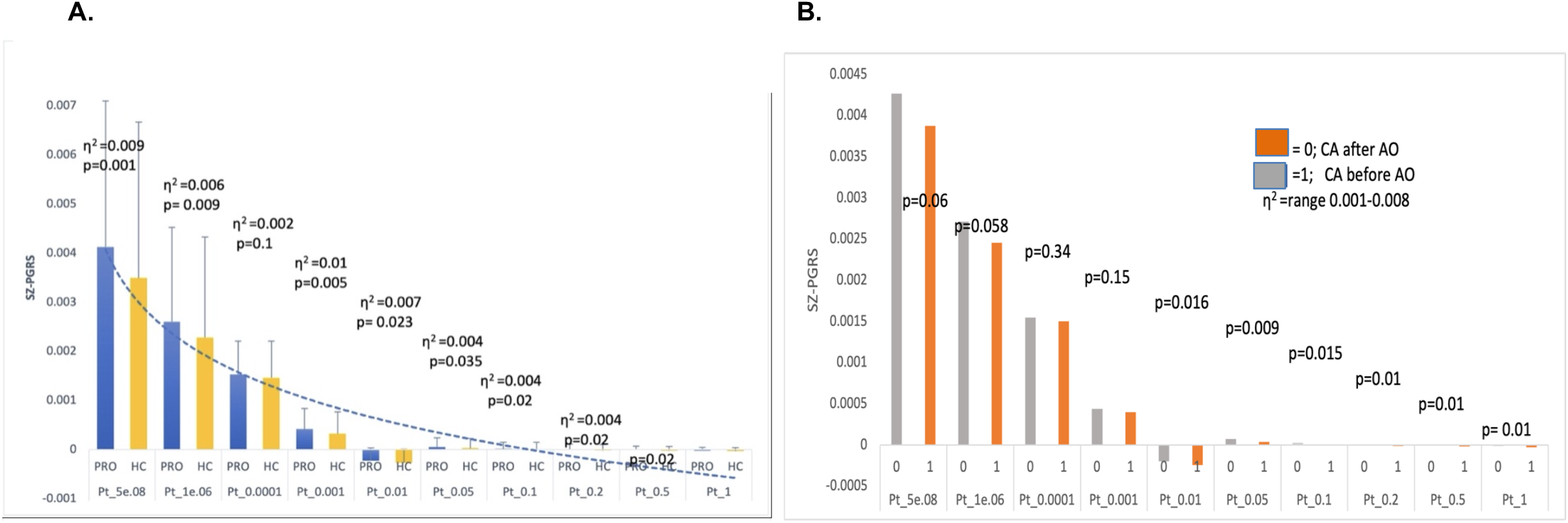
A. SZ-PGRS is higher in PRO compared to HC at all levels of risk SZ variants included. SZ-PGRS was corrected for sex, age, ancestry. In blue, PRO, in yellow, HC. The η^2^ represent the effect size and the amount of variance the PGRS contributes to group differences in the BSNIP2 dataset. B. SZ-PGRS is higher in probands with CA usage before PSY Onset.

**Supplementary Table 1.**
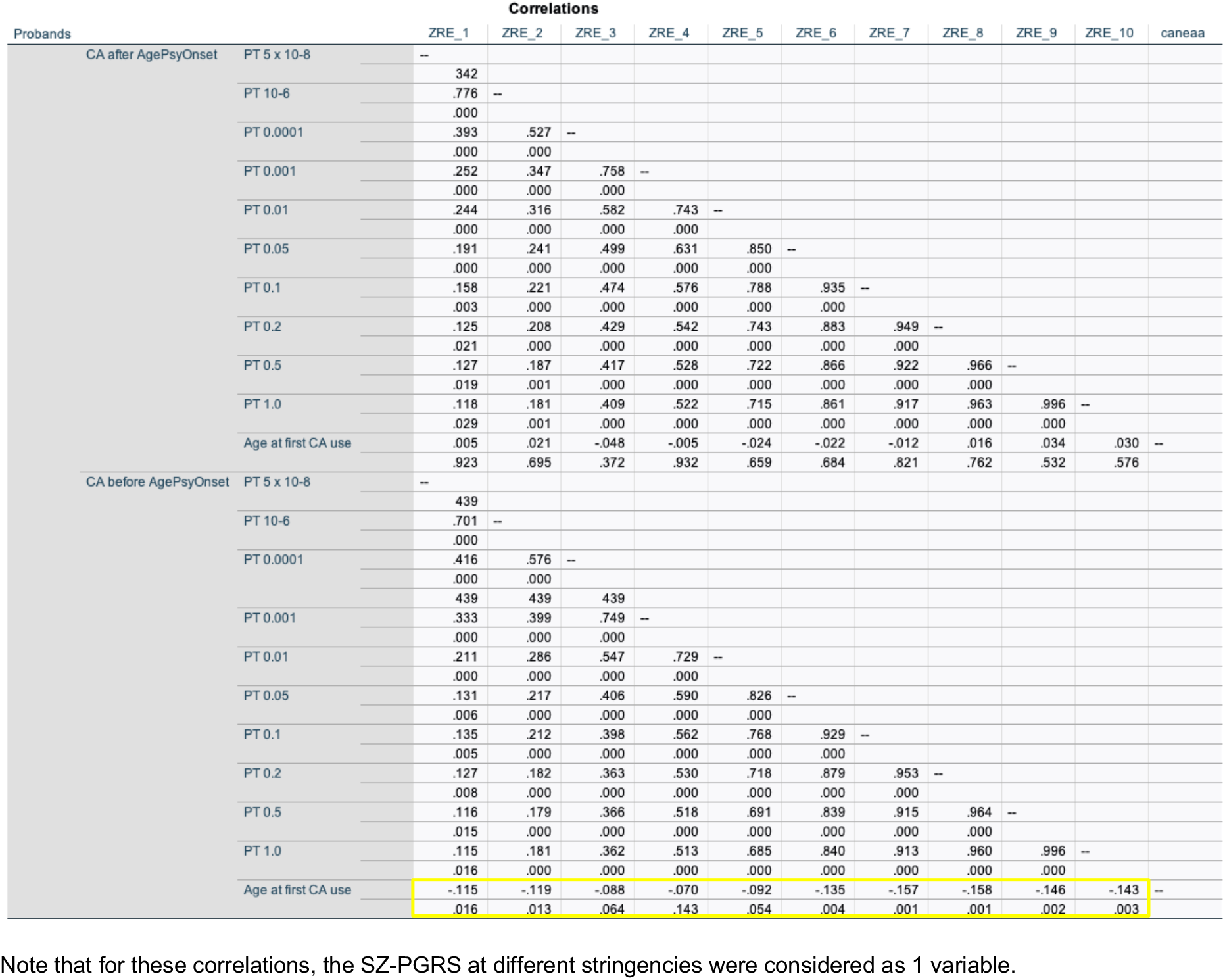
Correlation between SZ-PGRS at 10 stringencies values and AgeCAOnset first use, in probands with CA use before or after AgePsyOnset.

**Supplementary Table 2.** Number of cases included in survival analysis

| **Case Processing Summary** |  |  |  |  |  |
| --- | --- | --- | --- | --- | --- |
| CT Strata | CA Strata | Total<br>N | N of<br>Events | Censored<br>N | Percent |
| low | none | 336 | 76 | 260 | 77.4% |
|  | recreational | 212 | 144 | 68 | 32.1% |
|  | abuse | 25 | 25 | 0 | 0.0% |
|  | dependence | 52 | 52 | 0 | 0.0% |
|  | Overall | 625 | 297 | 328 | 52.5% |
| moderate | none | 92 | 48 | 44 | 47.8% |
|  | recreational | 162 | 148 | 14 | 8.6% |
|  | abuse | 22 | 22 | 0 | 0.0% |
|  | dependence | 57 | 57 | 0 | 0.0% |
|  | Overall | 333 | 275 | 58 | 17.4% |
| medium | none | 34 | 26 | 8 | 23.5% |
|  | recreational | 73 | 72 | 1 | 1.4% |
|  | abuse | 14 | 14 | 0 | 0.0% |
|  | dependence | 23 | 23 | 0 | 0.0% |
|  | Overall | 144 | 135 | 9 | 6.3% |
| high | none | 15 | 13 | 2 | 13.3% |
|  | recreational | 33 | 33 | 0 | 0.0% |
|  | abuse | 4 | 4 | 0 | 0.0% |
|  | dependence | 11 | 11 | 0 | 0.0% |
|  | Overall | 63 | 61 | 2 | 3.2% |
| Overall | Overall | 1165 | 768 | 397 | 34.1% |

### Participants Inclusion/exclusion criteria

Inclusion criteria for all participants included males and females aged 18-60, as well as proficiency in English. Participants in the probands group were assigned a DSM-IV-TR diagnosis based on interviews by trained clinical interviewers and must have met DSM criteria for SZ, SAD, or BP. HCs must have had no personal history of any psychotic or bipolar disorders, recurrent major depressive disorder, or a family history of SZ, SAD, or BP in first degree relatives. Exclusion criteria for all participants included: an estimated premorbid IQ < 60; major neurological or cognitive disorder (e.g., seizure disorder, traumatic brain injury, cerebrovascular disease, pervasive developmental disorder, mental retardation); serious medical, neuro-opthalmological, or neurological illness that could affect CNS functioning (e.g., decompensated cardiovascular disease, decompensated chronic obstructive pulmonary disease, late stages of diabetes, AIDS); women who are pregnant or breastfeeding (due to unknown risks related to MRI exposure) or a positive urine pregnancy test on day of MRI; and, presence of medical (e.g., artificial joints, brain aneurism clips, surgical pins, rods, wires, implants) or non-medical (e.g., metal piercing) irremovable metallic objects on/inside body (due to MRI-relevant risks). Additional exclusion criteria included recent substance use disorders (dependence within the past 3 months or abuse within the past 1 month) or persistent drug dependence lasting > 2 years in the previous 5 years to exclude cases of substance-induced psychosis or evidence of known neurological sequelae of drug use. We also excluded participants with positive urine drug screens on the day of assessment.

### Participants received a comprehensive symptom and cognition battery

Participants were evaluated with the global assessment of function (GAF) and the composite score using the Brief Assessment of Cognition for Schizophrenia.^52^ BACS scores were pre-adjusted for age and sex, z-transformed and then winsorized to +/- 3.0 standard deviations. Daily dose chlorpromazine equivalent (CPZ) were calculated for those taking antipsychotics. Childhood trauma data were acquired by supervised self-reported Childhood Trauma Questionnaire (CTQ) (Bernstein 1994). The age of psychotic symptoms onset (AgePsyOnset) as well as CA use data were acquired by trained clinicians using a semi-structured manner, as well as the SCID interviews. AgePsyOnset was determined as the first appearance of psychosis symptomatology. CA use was categorized as ever use, yes or no; and amount of usage, recreational, defined as seldomly in the past without meeting criteria for abuse or dependence; abuse and dependence according to the definition of the DSM IV (American Psychiatric Association 1994). We assessed CA of participants at any time, as well as when CA was initiated before or after AgePsyOnset (For some subjects age at initiation was not available). For all ratings, clinical raters were asked to rate confidence in self-reporting measures on a scale of 1-3, with 1 being very confident. The mean score for confidence was 1.29 ± 0.48, and a median of 1.0.

### Genotypes

Genotypes pre-imputation QC included missingness by marker <5%, missingness by sample <2%, minor allele frequency >1%, Inbreeding Coefficient (−0.2 > F_Het > 0.2), exclusion of monomorphic markers, Sex Check using X-chromosome heterozygosity and Y- chromosome call rate (all samples with sex mismatch were dropped). Detection of cryptic relatedness 2^nd^ degree or closer was done with KING (5). Imputation to 1000 Genomes Phase3 V5 panel (www.internationalgenome.org) was performed at the Michigan Imputation Server using programs Minimac4 (3) for imputation and Eagle (4) for phasing. Post-imputation QC included missingness by individual 5%, missingness by marker 1%, HWE P<1E-05, and MAF 1%. The final imputed dataset contained 10,321,126 SNP markers.

### SZ-PGRS

SZ-PGRS were generated with PRSice (www.prsice.info) using the Psychiatric Genomic Consortium Schizophrenia 2 (PGC-SZ2) training dataset, ^53^ for P thresholds 5e-8, 1e-6, 1e-4, 1e-3, 0.01, 0.05, 0.1, 0.2, 0.5 and 1. For group comparisons, the SZ-PGRS was corrected for the first two PCAs, age and sex.

